# Assessment of Airborne Disease Transmission Risk and Energy Impact of HVAC Mitigation Strategies

**DOI:** 10.1101/2021.11.15.21266233

**Authors:** Michael J. Risbeck, Martin Z. Bazant, Zhanhong Jiang, Young M. Lee, Kirk H. Drees, Jonathan D. Douglas

**Affiliations:** Johnson Controls International plc, Milwaukee, WI, USA; Departments of Chemical Engineering and Mathematics, Massachusetts Institute of Technology, Cambridge, MA, USA

## Abstract

The COVID-19 pandemic has focused renewed attention on the ways in which building HVAC systems may be operated to mitigate the risk of airborne disease transmission. The most common suggestion is to increase outdoor-air ventilation rates so as to dilute the concentrations of infectious aerosol particles indoors. Although this strategy does reduce the likelihood of disease spread, it is often much more costly than other strategies that provide equivalent particle removal or deactivation. To address this tradeoff and arrive at practical recommendations, we explain how different mitigation strategies can be expressed in terms of equivalent outdoor air (EOA) to provide a common basis for energy analysis. We then show the effects of each strategy on EOA delivery and energy cost in simulations of realistic buildings in a variety of climates. Key findings are that in-duct filtration is often the most efficient mitigation strategy, while significant risk reduction generally requires increasing total airflow to the system, either through adjusted HVAC setpoints or standalone disinfection devices.

## 1. Introduction

Throughout the past few decades, there has been significant emphasis on enhancing building HVAC systems to be more energy-efficient. In many cases, these measures include reducing ventilation rates and overall airflows to achieve corresponding energy reduction. Recently, the global COVID-19 pandemic has led to a significant reassessment of building operation, focusing on how HVAC systems may help to reduce the risk of airborne transmission of respiratory diseases. This new goal of infection risk mitigation often leads to the opposite recommendation, that outdoor-air ventilation be *increased* [1], to the detriment of energy efficiency [2, 3].

To assist HVAC practitioners in understanding the fundamental physics behind disease transmission and to manage the complexity of possible infection mitigation strategies, we discuss in this paper how the overall analysis can be framed in terms of equivalent outdoor air (EOA) delivery, as a basis for infection-risk and energy-consumption analysis. We then present the effects of possible HVAC mitigation strategies, highlighting cases that are particularly efficient or inefficient. Overall, we would like to emphasize the following key points:

- Increased outdoor-air ventilation is effective for reducing infection risk, but it can be very costly or even infeasible depending on climate and equipment configuration.
- Improved filtration is often the most cost-effective source of equivalent outdoor air.
- Significant reduction in infection risk generally requires increasing the total airflow supplied to zones, which may or may not be feasible depending on system configuration and ambient conditions.
- In cases where operational flexibility of the HVAC system is limited, high-risk zones may need supplementary in-zone filtration or other standalone disinfection devices.

To help frame our discussion, Fig. 1 gives an illustration of infectious particle dynamics and energy consumption associated with a typical HVAC system. In this scenario, the infector releases infectious aerosols (shown in red) into the air when they exhale. These particles quickly mix with the surrounding air where they undergo a variety of removal processes (shown in blue). In the meantime, the susceptible individual can potentially become infected if they are exposed to a high enough dose of the particles. To reduce this likelihood, the HVAC system can be adjusted or augmented so as to increase the removal rate of particles, thus reducing their average concentration. However, these operational measures almost always require an increase in energy consumption and in some cases can compromise occupant comfort. Thus, when choosing infection mitigation strategies, it is important to consider which strategies (or combination thereof) are most efficient.

**Figure 1:**
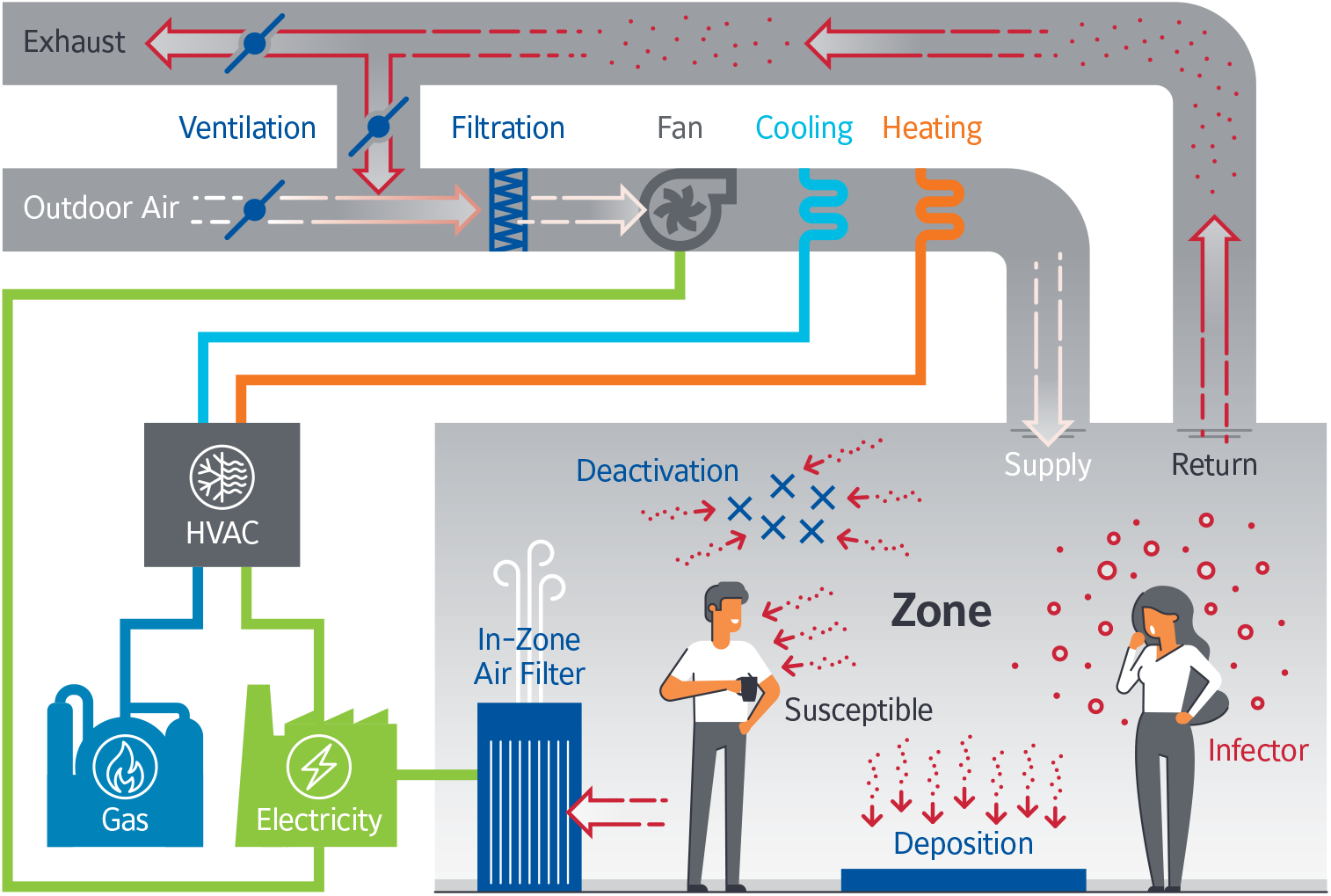
Diagram of infectious particle dynamics and HVAC energy consumption in typical buildings. Particle removal sources are shown in blue, while energy consumption is shown in purple.

## 2. Infection Risk Modeling and Equivalent Outdoor Air

It has long been understood that respiratory diseases, such as measles, tuberculosis, seasonal flu, and severe acute respiratory syndrome (SARS), can be transmitted through the air via exhaled aerosols (ranging from 0.1 to 10 µm in diameter) released into the air by infectious individuals. Indeed, it is now widely accepted that this is the dominant mode of transmission for COVID-19 [4, 5]. Generation and distribution of respiratory aerosols has been discussed in previous articles [6, 7], so in the interest of brevity, we will not review them in detail here.

Quantitative analysis of aerosol-based disease transmission dates back to Wells [8] and Riley et al. [9], who proposed that the infectious aerosols could be modeled as a standard indoor air pollutant. This approach allows steady-state concentrations to be estimated by finding the balance point between particle generation and removal. A key assumption of Wells-Riley type models is that the air within the space is *well-mixed*, i.e., uniform in particle concentration, which typically results from natural and forced turbulent convection associated with human respiration, temperature variations, and ventilation flows [10, 11]. Although the models do not account for short-range transmission via jet-like respiratory flows that can occur when infectious and susceptible individuals are in close proximity [12, 13], the models can also be corrected to account for the associated elevated risk [4].

Within the well-mixed model, a key observation is that the infectious particles can be effectively removed from the air via a variety of processes as follows:

- Ventilation: indoor air is vented and replaced with clean air from outdoors that is free from infectious particles.
- Filtration: recirculated indoor air passes through a filter, which traps some fraction of the infectious particles.
- Deactivation: through either natural decay or active measures (e.g., UVGI [14, 15]), the infectious material within the particles is destroyed; although the particles remain in the air, they are no longer infectious.
- Deposition: aerosols naturally deposit onto surfaces, which removes them from the air.

Note that for our purposes, we use “ventilation” to refer specifically to outdoor air, as the cleaning effect on recirculated air provided by the in-duct filter is captured in the “fitration” category. To compare and combine these effects, ASHRAE [16] has defined the concept of equivalent outdoor air (EOA), which gives the volumetric flow of outdoor air that would provide an equivalent removal rate of infectious particles. If particle size distributions are known, it is straightforward to determine the EOA provided by each of the removal mechanisms [2].

After the total EOA has been quantified, the model can be used to estimate the number of disease transmissions as

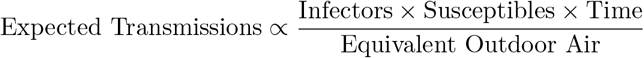

where the proportionality constant depends on a number of additional factors related to occupants (such as activity level, vocalization, immunity, viral strain, and whether masks are worn) and can vary by multiple orders of magnitude across different indoor spaces and occupancy characteristics [4]. For our purposes, it is enough to recognize that these factors are constant regardless of the operation of the HVAC system, and thus overall infection risk is inversely proportional to the EOA delivery rate.

## 3. Mitigation Strategies

Given that increased EOA delivery can reduce the risk of airborne disease transmission, it is desirable for system operators to do just that. Unfortunately, this cannot always be *directly* accomplished. For deposition and natural deactivation, these rates are essentially determined by the size distribution and biological response of the infectious aerosols; although there are weak dependencies on temperature and relative humidity [4], they are not strong enough to motivate changes to temperature or humidity setpoints in buildings. By contrast, the rates of ventilation and in-duct filtration can vary significantly, but they are essentially dictated by the operation of the HVAC system (and the in-duct filter type, in the case of filtration). Finally, active disinfection and in-zone filtration can be provided by portable devices, but of course these devices have to first be installed in the space.

Given these considerations, we identify four main decision variables relevant to EOA delivery in typical commercial AHU/VAV systems:

- AHU Minimum Outdoor Flow Setpoint: outdoor air provides a direct source of infectious-particle removal, although adjusting the *minimum* can have little to no effect during economizer conditions.
- AHU Supply Temperature Setpoint: raising the supply temperature setpoint increases the *total* airflow required to cool zones, thus providing additional filtration for recirculated air and possibly also additional ventilation during economizer conditions.
- In-Duct Filter Type: using a higher-efficiency filter results in more particles being removed from the recirculated air; however, if there is no recirculated air, then the filter does not provide any EOA.
- Activation of In-Zone Disinfection Devices: portable air filters or upper-room UVGI lamps can directly remove or deactivate infectious particles from zones independently of the operation of the HVAC system.

Note that in different systems, there may be different variables that can be specified (e.g., demand-controlled ventilation setpoint or economizer suitability temperature), but we select these variables as a representative cross section.

A key observation is that the first three variables listed above all operate on the same supply air stream, and thus they cannot always provide an independent source of EOA. For example, if the minimum outdoor flow setpoint is high enough that the system is operating near 100% outdoor air, then the in-duct filter essentially provides zero EOA, as there are no infectious particles for the filter to remove. The maximum possible EOA that can be delivered by these variables is the total supply flow provided by the HVAC system, which is tightly coupled to the thermal loads experienced by the space and thus often cannot be controlled directly. In addition, we note that the first two variables, if poorly chosen, can compromise occupant comfort. When outdoor-air conditions are particularly hot or cold, the system may not be able to provide the heating or cooling required to adequately condition additional outdoor air. Similarly, if the supply temperature setpoint is adjusted too far from its design value, the system may hit flow constraints without being able to meet the thermal loads in the space. Alternatively, setting the supply temperature setpoint too high may not provide sufficient dehumidification, leading to excessive zone humidity. These concerns are less relevant during milder shoulder seasons, but it is important to recognize that the HVAC setpoints may not provide as much flexibility as expected, and they may need to be adjusted gradually to ensure comfort is maintained.

Finally, we note that the selected variables are primarily applicable to commercial buildings with central HVAC systems. Therefore, the conclusions that follow may not apply to older buildings that are naturally ventilated or otherwise lack central HVAC. For example, in classrooms without mechanical ventilation, none of the indicated HVAC setpoints are available to be manipulated. While the use of stand-alone in-zone EOA sources is possible, such devices may be costly to deploy or lead to unacceptable noise levels. In such situations, the only readily available way to increase EOA delivery is to manually open windows [17]. Unfortunately, this strategy may compromise energy efficiency (and possibly also occupant comfort), but it may nevertheless be necessary to achieve acceptable risk levels. Thus, although the findings in this paper apply to a large class of buildings, the appropriate course of action will of course need to be modified for spaces with different HVAC capabilities.

## 4. Energy Analysis

Although the mitigation strategies can provide infection risk reduction via increased EOA delivery, employing these strategies almost always leads to an increase in overall energy consumption. We summarize this energy impact as follows:

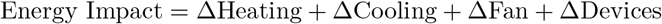

When increasing the minimum ventilation rate, the primary change to energy consumption is to heating and cooling for the outdoor air, and the magnitude of these changes varies significantly with outdoor-air conditions. It is possible that energy use may *decrease* in some conditions, but a well-functioning economizer would likely already capture these savings in the baseline case. Fan power may also increase if the change increases total flow to the zone. For an increase to the supply temperature setpoint, energy analysis is more complicated. Whenever this change leads to additional supply flow, fan power is increased. In the heating season, heating energy consumption may increase, although the the effect will depend on equipment configuration and sequence of operation. However, in the cooling season, the cooling energy consumption is likely to decrease, as the *latent load* required to meet the supply temperature setpoint is now smaller. Because the fan and cooling effects are opposite, the net overall effect may be positive or negative depending on which dominates. When changing the in-duct filter type, the only energy impact is on fan power, as different filter grades impose different pressure drops that must be overcome by the fan to meet airflow requirements. For standard MERV filters, the change in pressure drop is often small, which makes high-efficiency in-duct filtration an extremely cost effective source of EOA [18].

Finally, activation of in-zone devices requires additional electricity consumption. In many cases, these devices must be on or off, so the only required data is power consumption when active. However, the energy intensity of different device can vary significantly, with common residential devices requiring 0.2 to 1 W*/*cfm of total flow [19, Table 3], and similar variation for larger devices that would be more applicable in commercial settings. Variable-speed devices may have more complicated power characteristics, which presents a further opportunity for optimization. We note that when comparing in-zone devices it is important to focus on total EOA delivery and power consumption, rather than other metrics like filtration efficiency. Indeed, HEPA and similar filters impose significantly larger pressure drops than MERV filters [18]. Thus, a device using a high-efficiency MERV filter can often deliver the same EOA as a HEPA device but with significantly lower power consumption. Similarly, UVGI devices in ducts [14] or free-standing units [15] can almost completely deactivate infectious particles, but if total flow through the device is small, or carries a low concentration of infectious aerosols, the overall impact on infection risk will be correspondingly small.

Overall, the main observation is that the energy impact of ventilation and supply temperature changes is very tightly coupled to outdoor-air conditions, whereas the cost of higher-efficiency filtration or in-zone devices is relatively constant. Thus, when choosing a particular strategy, it is important to consider how long the measures are likely to be employed and what the weather conditions will be over that time period. Given these complexities, there is an opportunity to *optimize* the chosen strategy on a regular basis using relevant predictive models [3]. Otherwise, the best course of action is to choose a robust strategy that can be applied consistently and explore other options only when necessary.

## 5. Annual Trends

To illustrate the key considerations for EOA delivery and associated energy consumption, we present example cases of how these quantities vary throughout a typical year for a typical office building in various climates. For this purpose, we use EnergyPlus [20] simulations of the same “Large Office” reference building and apply each of the discussed infection mitigation strategies. Information about the chosen climate zones, along with key HVAC parameters, is shown in Table 1. Note that the HVAC system’s design flow varies slightly across the cases, as HVAC equipment is automatically sized by EnergyPlus based on the different design climates for each. Additional details are given in Appendix A.

**Table 1:**
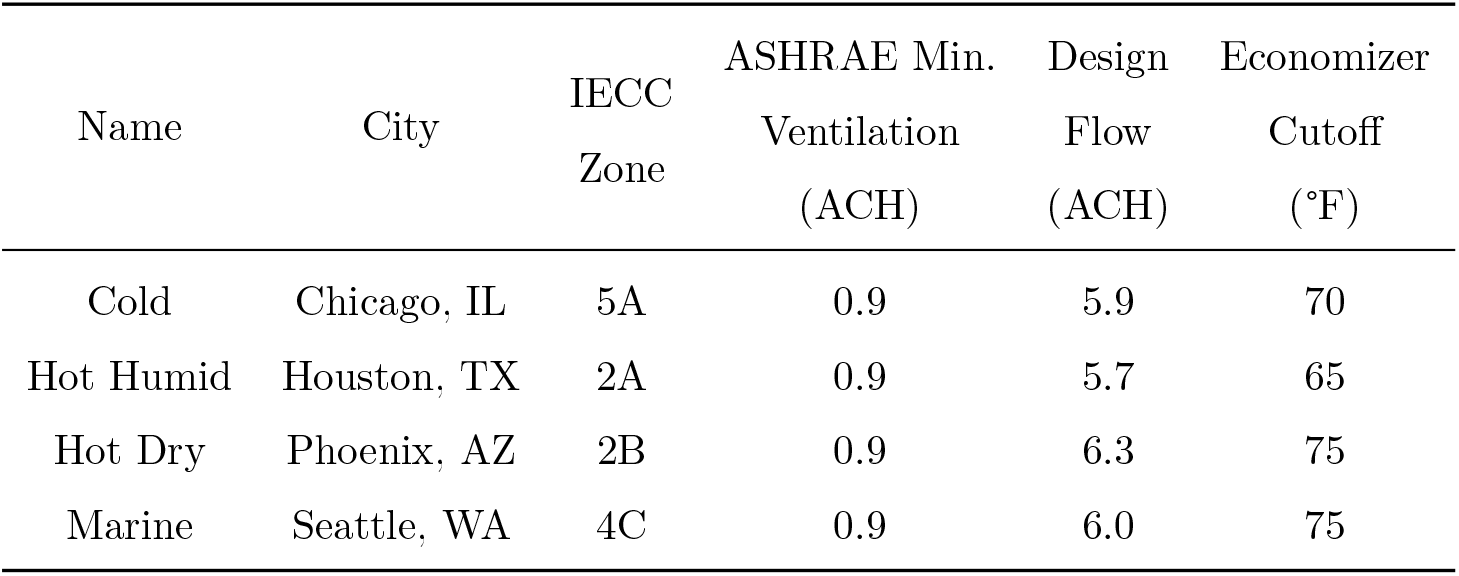
Information for example climate zones. Minimum ventilation is chosen by EnergyPlus in accordance with ASHRAE Standard 62.1 [21]. Economizer cutoff temperature is taken from ASHRAE Standard 90.1 [22].

For our purposes, we consider the baseline case to be the ASHRAE-specified minimum ventilation rate with a MERV8 in-duct filter and a supply temperature setpoint of 55 °F, which we refer to as the “ASHRAE Baseline”. From here, we then consider five different alternative cases:

- 0.5*×* Min. Ventilation: minimum ventilation rate set to half the ASHRAE value (to represent buildings that are not adequately ventilated)
- 2*×* Min. Ventilation: minimum ventilation rate set to double the ASHRAE value
- MERV13 Filter: in-duct filter switched from MERV8 to MERV13
- Higher Supply Temperature: supply temperature setpoint increased from 55 °F to 62 °F
- +1 ACH In-Zone Filtration: add portable HEPA filtration units to provide an extra 1 ACH of EOA (device power 0.65 W*/*cfm)

These cases represent the possible mitigation strategies discussed in Section 3. For cost calculation purposes, we assume cooling is provided with a COP of 3 and heating with an efficiency of 90% for electricity at 0.12 $*/*kWh and gas at 8 $*/*MMBTU.

### 5.1. EOA Breakdown

To gain insight into how each of the mitigation strategies provide additional EOA, we provide a weekly breakdown of EOA delivery by source for each climate and case in Fig. 2. All values are averaged over occupied hours for each week. Note that both the “Economizer” and “Min. Ventilation” categories are due to ventilation, with the former indicating discretionary extra ventilation above the minimum provided by the economizer (which varies with outdoor-air temperature). For reference, these figures also show the change in *total* energy cost throughout the year relative to the baseline case. Note that for brevity we omit the plots for the in-zone filtration cases, as it simply shifts the baseline case up by 1 ACH of EOA. In each set of axes, the black “Max EOA” line represents the maximum possible EOA delivery for that system assuming 100% clean supply air. These conditions would be achieved under either 100% outdoor-air operation or using an in-duct filter with 100% capture efficiency. Although system capacities are generally near 6 ACH, such high flows are only provided on the hottest hours of the hottest days, and thus the weekly averages shown in the figure are always well below that value.

**Figure 2:**
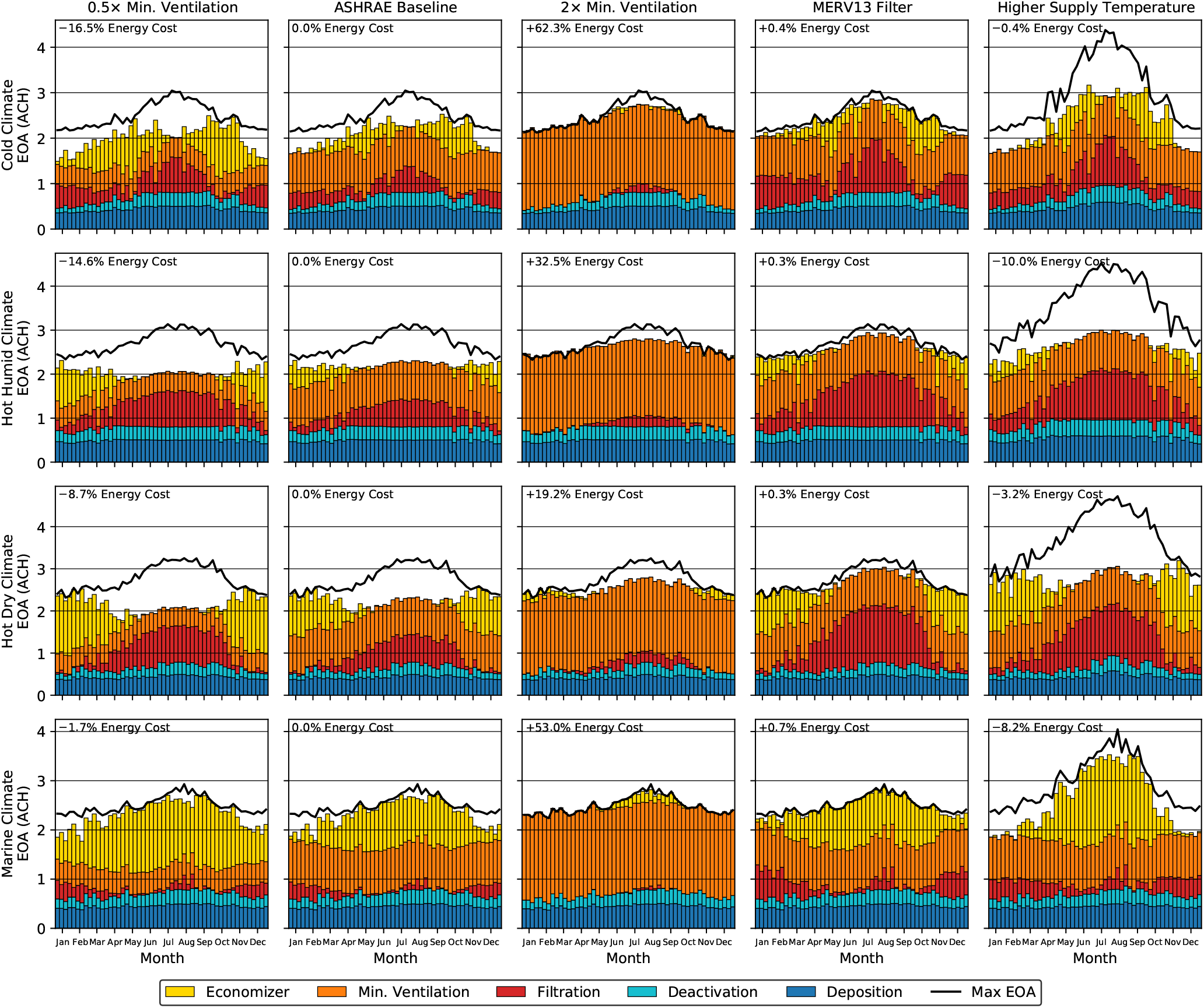
Weekly breakdown of EOA sources for an office building in various climates. Values are averaged over the occupied period for each week in one year of operation. Labeled energy costs are relative to baseline operation averaged over the whole year.

Starting in the upper-left corner of Fig. 2 and moving right, we note that increasing from half to the full ASHRAE-required ventilation rate often does not have a significant impact on EOA delivery but does noticeably increase energy costs. The explanation for this behavior is that under economizer conditions, the minimum ventilation rate does not matter (because the system is operating at 100% outdoor air), and otherwise the in-duct filter is able to partially clean the extra recirculating air (roughly 40% of it, in the case of the MERV8 filter) that would otherwise come from outside. Thus, as the orange bars increase, the red and yellow bars also shrink. Moving further to the right, we see that doubling the minimum ventilation rate from ASHRAE requirements does provide increased EOA, but not as much as might be expected. In this case, the reason is exactly the same as before: the increase in the “Min. Ventilation” category comes with a corresponding decrease in the “Economizer” and “Filtration” categories. Note that at this ventilation rate, the system is at 100% outdoor air for nearly the entire year, and thus EOA delivery is near its maximum value. However, moving to the fourth column, we see that similar EOA delivery can be achieved by leaving the ventilation rate at its ASHRAE value and instead switching to a MERV13 filter. Finally, moving to the last column we see the most interesting results associated with the higher supply temperature. In this case, whenever the HVAC system is operating above minimum flow constraints, the total supply airflow is increased by the control layer as required to maintain temperature setpoint. Thus, both the maximum possible and actual EOA delivery rates increase relative to the ASHRAE baseline case.

Moving down to the next two rows associated with Hot climates, the observed trends are similar but with different annual distribution due to the relevant seasons. In these climates, we note that the economizer season is shifted primarily from Spring and Fall months to the Winter. Thus, the increased energy costs associated with ventilation are almost exclusively due to cooling, in contrast to the mixture of heating and cooling that affected the Cold climate. We note also that the total cost penalty for the Hot Dry climate is less than that of the Hot Humid climate because the dry climate does not see the significant latent cooling load observed for the humid climate. As before, we see that the higher supply temperature case in column four is the only case that increases total supply flow, but unfortunately not all of this potential EOA can be realized due to operating at minimum-ventilation conditions. By contrast, moving to the Marine climate in row four, we see that economizer operation means the system is operating near 100% outdoor air throughout much of the year, and thus near maximum EOA is achieved regardless of the operating strategy. (We do note that the large increase in energy cost for the 2*×* ventilation case is primarily due to heating costs, as will be discussed in the next section.)

Overall, these results illustrate that due to the coupling of ventilation and in-duct filtration, there are diminishing returns for each strategy, and the associated maximum EOA delivery is limited by the *total* supply air flow provided by the HVAC system. Because none of the variables in the first four columns change supply flow, they all have the same upper bound. By contrast, the higher supply temperature setpoint in the final column *does* increase total supply flow and thus also the maximum possible EOA delivery. Indeed, we note that combining the higher supply temperature with a MERV13 filter would provide considerably more EOA compared to the baseline case (almost the maximum value indicated by the black line). However, some care is needed when adjusting supply temperature to ensure that occupant comfort is not sacrificed. The primary concern is that a higher supply temperature will provide inadequate dehumidification. Although in this simulation, weekly average humidities for all climates remained below 65%, spaces with higher occupant densities and higher moisture loads may not be able to tolerate 62 °F supply air. (For this reason, ASHRAE Guideline 36 [23] suggests an upper limit of 65 °F in mild or dry climates and 60 °F or lower in humid climates.) Thus, while raising the supply temperature setpoint is an intriguing possibility for many spaces, the best widely applicable solution for EOA delivery is to improve in-duct filtration to MERV13 or better.

### 5.2. Energy Costs

Consistent with the EOA observations in the previous section, we turn now to the associated energy consumption. For this purpose, we show monthly average EOA and energy cost for each climate and case in Fig. 3. In addition, to determine which of the EOA sources are most energy-efficient, we plot a monthly “price” of the additional EOA as the ratio between the change in energy cost and change in EOA delivery from the baseline case. This value essentially normalizes all strategies to providing an additional 1 ACH of EOA and then estimates the energy impact. In general, we prefer strategies with a low (and ideally negative) EOA price.

**Figure 3:**
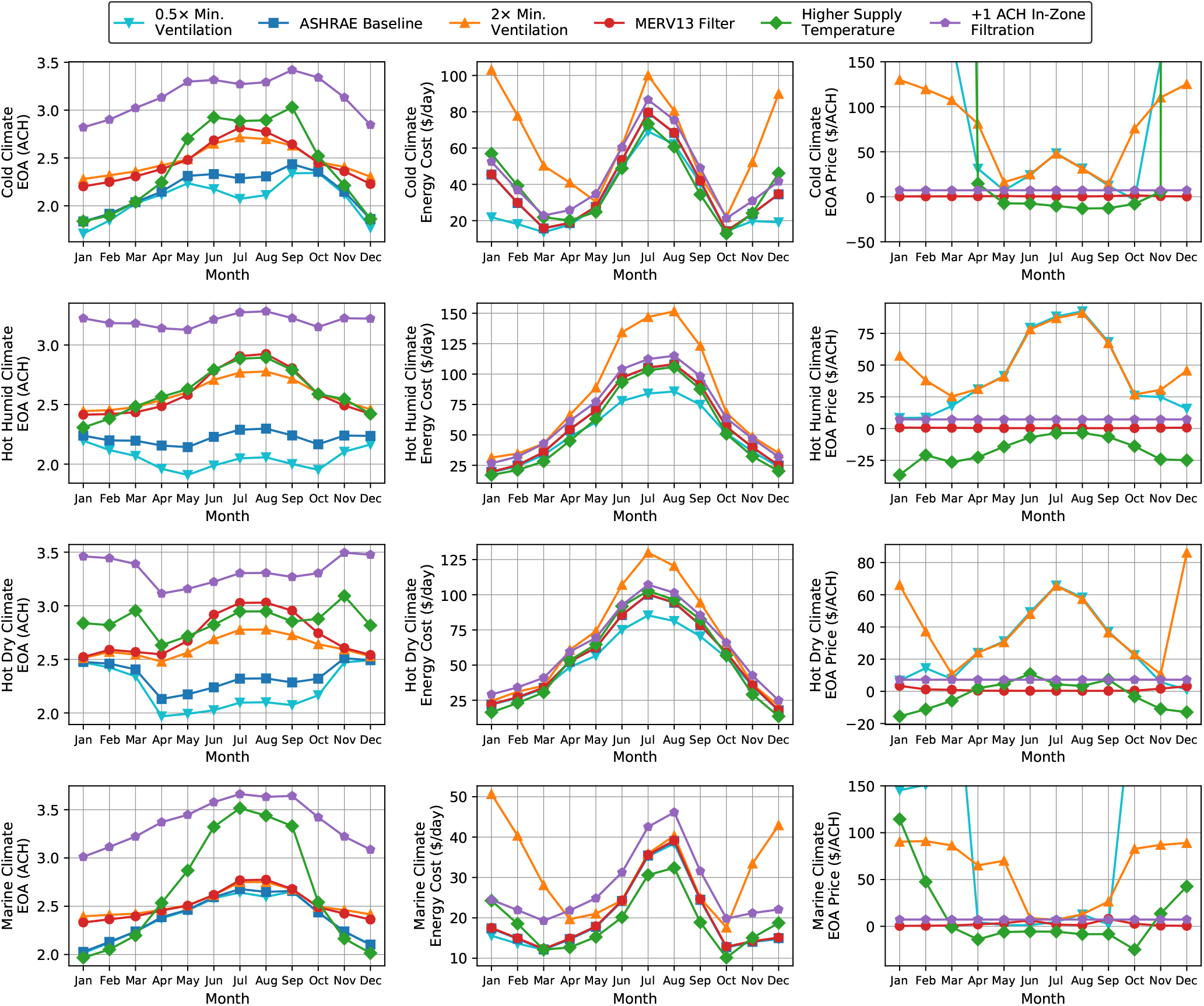
Annual trends in EOA and energy cost for baseline operation and various infection mitigation strategies in each climate. First two columns show monthly averages for EOA and energy cost, while last column shows ratio of change in EOA to change in energy cost relative to baseline operation.

Starting in the first column of Fig. 3, we note that the EOA delivery are the same values as in Fig. 2 but re-averaged on a monthly basis and with a new curve for the in-zone filtration case. With the values overlaid, it is much easier to see which strategies are providing the most EOA. We see here that in-zone filtration is the only consistent strategy throughout the year, as those devices can be deployed at will independent of the HVAC system. Both the increased ventilation rate and the MERV13 filter also provide additional EOA each month, but the magnitude varies throughout the year. Finally, we see that the higher supply temperature is the most inconsistent source of EOA, generally providing no additional EOA during heating conditions but significantly more during cooling conditions. Moving to the second column, the most obvious trend is that energy consumption is driven primarily by the annual climate cycle, with the disinfection strategies shifting costs only marginally from the baseline case. The obvious outlier to this trend, however, is ventilation during heating conditions. For the 0.5*×* ventilation case, internal building loads are often enough to meet the heating requirements of the outdoor air, and thus energy consumption is quite low. However, as the outdoor air rate is increased to the ASHRAE standard, some supplemental heating is required, and at 2*×* ventilation, a very large amount of heating is needed.

To examine the primary tradeoff between EOA delivery and energy cost, we move now to the third column in Fig. 3, which estimates the per-ACH price of EOA delivered by each scenario. Because this quantity is calculated as a ratio, a strategy can have a high price if it requires significant additional energy consumption (i.e., a large numerator) but also if it delivers little to no supplemental EOA (i.e., a small denominator). For the Cold climate in the first row, we see that both ventilation cases and the higher supply temperature case have very high EOA prices during the heating season, but during the cooling season they diverge, with the higher supply temperature actually having a slightly negative price (due to its reduction of latent cooling loads). By contrast, both the MERV13 filter and the in-zone filtration devices have nearly constant EOA prices throughout the year. Based on this metric, the MERV13 filter is the clear winner, as its effect on energy cost is nearly zero, but if the resulting EOA delivery is not sufficient, it could be combined with other sources. For the two Hot climates (in rows two and three), we note that there is no heating season, and thus the cooling-season trends apply throughout the year. Under these conditions, extra ventilation is particularly costly and should likely be avoided in favor of an alternative strategy. Once again, raising the supply temperature setpoint is a particularly attractive option based on this comparison, but some care is needed to ensure that the HVAC system still provides adequate dehumidification. Finally, the Marine climate in the last row is generally similar to the cold climate, although with significantly lower average energy cost.

The key observation from this analysis is that ventilation is generally a particularly costly source of EOA, and so, if a building is already receiving the ASHRAE minimum outdoor air (or if CO_2_ levels indicate acceptable air quality and transmission risk [24]), then other strategies should be examined to provide additional EOA and thus further reduce infection risk. In particular, upgrading to a MERV13 filter provides extremely cheap EOA and is almost equivalent to operating at 100% outdoor air due to its high filtration efficiency. After upgrading the in-duct filter, a higher supply temperature during the cooling season is worthy of exploration, but other system limitations may prevent the use of this strategy. In such cases, the most robust option for EOA delivery is the installation of in-zone filtration units, although capital and operating costs will depend on the particular devices that are selected.

## 6. Conclusions

In this paper, we have analyzed the dynamics of infectious aerosol particles in buildings, formulating their removal in terms of EOA. Based on our model, we have suggested possible actions that could be taken to increase EOA delivery (thus reducing infection risk) along with the corresponding effect on energy consumption. We then illustrated these considerations via building simulations across a range of climates, making the following key observations:

- Outdoor-air ventilation is a direct source of EOA, but it may already be provided by an economizer or made redundant by a high-efficiency filter, and thus it is often the most energy-inefficient source of EOA.
- Improved in-duct filtration efficiency provides additional EOA with only a small increase in energy cost, and thus it is generally the first infection mitigation action that should be taken.
- Raising the supply temperature setpoint is often the only direct way to increase total supply airflow and EOA while often reducing energy cost, but it may lead to unacceptably high indoor humidity or temperature due to reduced cooling capacity.
- Standalone filtration units or other disinfection devices are the only independent way to provide EOA, and they are often cost-competitive with HVAC sources.

We hope that these insights may help to guide operational strategies during the current COVID-19 pandemic and beyond. Our models may be extended to new viral strains or other respiratory infections by simply varying the relative susceptibility [4]. The analysis can also be applied more broadly to indoor air quality to help justify similar design and operational changes, which may be further informed by carbon dioxide monitoring [24].

## Data Availability

All data produced in the present work are contained in the manuscript.

## Appendix A. Simulation and Calculation Details

The examples show in Section 5 are generated using simulation data from Energyplus 9.4 [20]. Annual simulations are run using the `RefBldgLargeOfficeNew2004_Chicago.idf` provided with EnergyPlus. The HVAC configration is left at the default settings in that file, except that the outdoor air economizers are changed from type `DifferentialDryBulb` to `FixedDryBulb` using the cutoff temperatures shown in Table 1. For the four different climates, design-day weather information is updated to use appropriate values based on TMY data for the cities shown in Table 1. HVAC equipment capacities are thus chosen automatically by EnergyPlus based on this information using its default autosizing algorithms. To simulate alternative setpoints (minimum ventilation and supply temperature), separate annual simulations are run using the same files with the appropriate setpoints adjusted and held constant for the full simulation period. The simulation timestep was set to 15 minutes.

After running the EnergyPlus simulations, the necessary data streams are extracted from raw simulation output. The results shown in Section 5 all use data taken from the first floor of the building, which covers the five zones `CORE_BOTTOM`, `PERIMETER_BOT_ZN_1`, `PERIMETER_BOT_ZN_2`, `PERIMETER_BOT_ZN_3`, and `PERIMETER_BOT_ZN_4`, all of which are served by the `VAV 1` airloop. To calculate EOA delivery and energy consumption, the following values are required:

- Outdoor air flow
- Outdoor air temperature
- Outdoor air (relative) humidity
- Total supply air flow
- Supply air flow
- Supply air (relative) humidity
- Average zone temperature
- Average zone (relative) humidity

Zone values are averaged over the five individual zones weighted by time-varying supply air flow to each zone, falling back to floor-area weighting when total supply air flow is zero. Note that the flow values output from EnergyPlus are in mass-flow units but are converted to volumetric-flow units via the appropriate air density. Where appropriate, the flows were further converted to volume-independent units (i.e., ACH) using the total space volume across the five zones. EOA flows and energy consumption are then calculated from these values on a 15-minute basis and then averaged or summed over the appropriate days and times for presentation in Figs. 2 and 3.

Calculation of EOA flows is largely as presented in existing works [2, 3]. Specifically, EOA is calculated for each of the categories as follows:

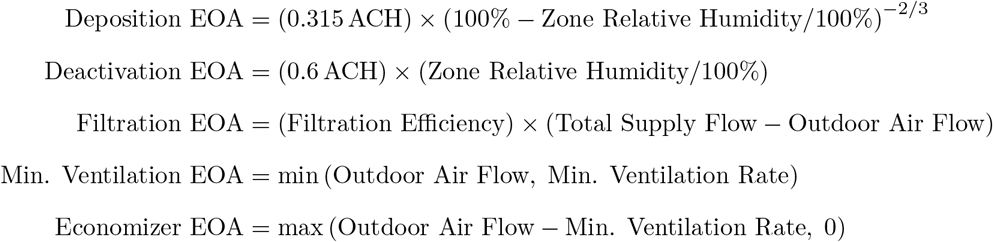

Deposition and deactivation follow the humidity dependence suggested in Bazant and Bush [4]. Filtration is calculated using only the *recirculated* portion of the supply air flow using an effective filtration efficiency that depends on the chosen filter type [16]. Ventilation is split into two components due to the required minimum ventilation and the additional discretionary ventilation provided by the economizer. Note that the first two categories are calculated directly in ACH units, while the last three categories must be converted from volumetric-flow units using the total zone volume. Results are averaged over the occupied hours for each day in the simulation period, with weekly results shown in Fig. 2 and monthly results in Fig. 3.

Energy consumption for AHU heating and cooling coils is calculated using standard thermodynamic relationships [2]. Cooling is assumed to be provided by electricity with a COP of 3, while heating is assumed to come from gas with an efficiency of 90%. Energy consumption for fans is calculated assuming a cubic dependence on flow based on the design flow and energy consumption from EnergyPlus. Energy consumption for in-zone devices is calculated based on device characteristics as described in the text. These four categories are then scaled by the appropriate utility prices (gas at 8 $*/*MMBTU for heating electricity at 0.12 $*/*kWh for the other three categories) and then summed to calculate total energy cost. Results are summed over each month in the simulation period and presented in Fig. 3.

